# Electronic prescribing systems as tools to improve patient care: a learning health systems approach to increase guideline concordant prescribing for venous thromboembolism prevention

**DOI:** 10.1101/2021.01.11.21249606

**Authors:** S. Gallier, A. Topham, P. Nightingale, M. Garrick, I. Woolhouse, M.A. Berry, T. Pankhurst, E. Sapey, S. Ball

## Abstract

**BACKGROUND:** Venous thromboembolism (VTE) causes significant mortality and morbidity in hospitalised patients. In England, reporting the percentage of patients with a completed VTE risk assessment is mandated, but this does not include whether that risk assessment resulted in appropriate prescribing. Full guideline compliance (an assessment and action) is rarely reported. Education, audit and feedback enhance guideline compliance but electronic prescribing systems(EPS) can mandate guideline-compliant actions. We hypothesised that EPS-based interventions would increase full VTE guideline compliance more than other interventions.

**METHODS:** All admitted patients within University Hospitals Birmingham NHS Foundation Trust were included for analysis between 2011-2020. The proportion of patients who received a fully compliant risk assessment and action was assessed over time. Interventions included face-to-face feedback based on measured performance (an individual approach) and mandatory risk assessment and prescribing rules into an EPS (a systems approach).

**RESULTS:** Data from all 235,005 admissions and all 5503 prescribers were included in the analysis. Face-to-face feedback improved full VTE guideline concordance from 70% to 77% (p=<0.001). Changes to the EPS to mandate assessment with prescribing rules increased full VTE compliance to 95% (p=<0.001). Further amendments to the EPS system to reduce erroneous VTE assessments slightly reduced full compliance to 92% (p<0.001), but this was then maintained including during changes to the low molecular weight heparin used for VTE prophylaxis.

**DISCUSSION/ CONCLUSION:** An EPS-systems approach was more effective in improving sustained guideline-compliant VTE prevention. Non-compliance was still not eradicated despite this mandated system and requires further research.

**FUNDING:** HDR-UK Hub PIONEER

**Summary Box:** *What is already known?:* Hospitalised patients are at an increased risk of venous thromboembolism (VTE), which can lead to significant morbidity and mortality. Risk factors for VTE are well known, there are established screening criteria and there is an effective prophylactic therapy, using low molecular weight heparin where indicated. Since 2010, NHS England has mandated the reporting of the percentage of patients with a completed VTE risk assessment. However, it does not automatically follow that completing a risk assessment leads to the appropriate action (prescribing and administering VTE prophylaxis where indicated). Currently it is unclear what percentage of patients have a guideline compliant VTE risk assessment and an associated action, or how full guideline compliance can be improved.

*What does this paper add?:* First, this paper describes that a VTE risk assessment does not always lead to full VTE guideline compliance (an appropriate prescription and administration of heparin where indicated). This is currently not part of mandated reporting but potentially could lead to patient harm. Second, that Electronic Health Systems (EHS) can capture and interrogate guideline-associated risk assessments and prescribing, and be used to improve full guideline compliance, through a combination of individual feedback to prescribing outliers and mandated prescribing rules. These EPS-assisted systems are robust, and sustain guideline compliance through personnel and formulary changes.

## Introduction

Hospital Acquired Thromboembolism (HAT) is defined as a venous thromboembolic event (VTE) (a deep vein thrombosis (DVT) and/or pulmonary embolus (PE)) which was not present on admission but diagnosed within hospital or within 90 days of hospital discharge^(1)^. HAT accounts for a significant amount of potentially avoidable morbidity and mortality^(2)^ with an incidence rate of 9.7 per 1000 hospital admissions, with 71% diagnosed post-discharge^(3)^.

The risk factors for VTE during admission are well known^(4)^, there is an effective prophylactic therapy^(2)^ and there are well-validated guidelines to assist assessment of the risk of VTE and where prophylaxis therapy should be prescribed^(5)^. However, rates of concordance with guidelines vary across international providers, from 16 to 85%^(6, 7)^.

In England, a Commissioning for Quality and Innovation (CQUIN) framework set a threshold rate for acute hospital providers to undertake risk assessments for at least 90% of inpatients each month in 2010-2011^(8)^, which increased to 95% by 2013^(9)^. This reporting is nationally mandated with each acute care hospital providing the number of adults admitted each month who have had a VTE risk assessment and the number of adults admitted in total, with specific groups of patients excluded from this analysis^(10)^. A recent National Health Service report suggests that the majority of acute hospitals in England are meeting the target of 95% of patients having a VTE risk assessment^(11)^. However, there is a difference in completing a risk assessment and acting upon it (either prescribing low molecular weight heparin (LMWH) when it is indicated or not when it is not). Currently, there are no national figures to describe or targets to ensure full VTE guideline compliance (assessment and action on assessment).

Prescribing errors include inappropriate or erroneous inclusion or omission of drugs and an inappropriate assessment of the potential harm from giving or omitting a treatment^(12)^. Not acting on a VTE risk assessment is a prescribing error. The potential reasons for prescribing errors are complex but studies suggest that most mistakes were made because of slips in attention, or because prescribers did not apply relevant rules^(13)^. A number of reports have highlighted strategies to improve prescribing practices^(14)^ including education, prescription aids (both paper and computerised)^(15)^, mandated prescribing^(16)^ and prescriber feedback and audit^(17)^. Given the relative short-term placements of junior medical staff within a specific hospital, targeting individual prescribing may be less effective for a single centre than implementing systems change to enhance safety^(18)^.

Electronic prescribing systems (EPS) have been shown to improve inpatient medication management, especially by reducing medicine-reconciliation, dose, and avoidable delay-of-treatment errors ^(18, 19)^. It is less clear whether EPS can reduce venous thromboembolic (VTE) prescribing errors associated with guideline non-compliance^(20)^ and whether this is sustained or can be improved with further support in the prescribing process.

University Hospitals Birmingham NHS Foundation Trust (UHB) is one of the largest NHS Trusts in England, providing direct acute services and specialist care across four hospital sites. The analysis was conducted at Queen Elizabeth Hospital Birmingham where an in-house built, clinically-led EHR Prescribing Information and Communications System (PICS) is present. PICS is a rules-based prescription-support system that includes all clinical documentation for admission, physiological and laboratory measurements, provides real-time drug prescribing checks and recommendations by triangulating physiological and laboratory results, comorbidities and prescribing data, as well as supporting institutional and individual audit of prescribing practices.

UHB reports a VTE risk assessment completed in 98.5% of admitted patients for 2019-2020^(11)^ and has met all national targets since 2010. However, hospital staff wanted to ensure that these risk assessments were being acted upon, with full guideline compliance (a risk assessment and appropriate prescription and administration of LMWH when indicated, or no prescription of LMWH when not indicated).

Prior to the implementation of the VTE prophylaxis interventions described in the current paper, all staff received a hospital induction to ensure familiarity with procedures and policies. This included a specific induction talk with a focus on VTE assessments and the importance of prescribing LMWH. Additionally, there were rolling lectures held by Consultant Haematologists to reinforce learning. Online videos were available on prescribing LMWH along with other key clinical focus areas.

We hypothesised that systemic prescribing interventions would be more effective in improving full VTE guideline compliance, compared to face to face educational interventions, and that these improvements would be sustained.

Prescribing interventions were:

- Feedback of individual performance of doctors with identification and interviewing outliers (known as the Junior Doctor Clinical Dashboard)
- An EHR systems approach of adding mandatory VTE steps to the EHR during the admission processes.
- A systems approach change to the EHR VTE assessment form after clinician feedback to reduce options to “work around” mandated prescribing proposals.

## Methods

### Ethical approval

This research was conducted under the ethical approvals of PIONEER, a Health data research hub in acute care (East Midlands – Derby Research ethics committee, reference 20/EM/0158).

### Patients

All emergency and elective adult admissions to the Queen Elizabeth Hospital Birmingham (part of University Hospitals Birmingham NHS Foundation Trust) from January 2011 to October 2020 were included. Post first intervention, all data collection and analyses were prospective.

#### Prescribing practices outcome

The primary outcome for this study was full VTE guideline concordance within 24 hours, defined as a risk assessment completed and the recommended treatment prescribed, expressed as a proportion for each week. Guideline non-compliance was defined as no VTE assessment performed, or where the VTE risk score suggested prophylaxis was needed and yet it was not prescribed or VTE prophylaxis was not indicated, yet it was prescribed.

### Interventions

Interventions were developed in discussion with hospital management, pharmacists, hospital doctors of all grades and specialties and bio-informaticians. Interventions were designed after discussion with clinical specialists in that field and were compliant with national guidelines for patient care.

#### INTERVENTION 1 – EDUCATION BY INDIVIDUAL FEEDBACK: THE JUNIOR DOCTOR CLINICAL DASHBOARD

The Junior Doctor Clinical Dashboard was developed utilising z-scores. For VTE prophylaxis this was based on their correct assessment and prescription of VTE prophylaxis on the PICS against current guidelines^(5)^. Z-scores were calculated using the following method.

If doctor “*I*” correctly treats “r_i_” cases from “n_i_” cases the expected proportion p of correct responses across all doctors is 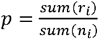 and has standard deviation 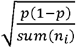. The observed proportion of correctly treated patients by doctor *i* is 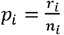 and has standard deviation 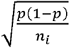. The standard deviation uses the expected proportion p of correct responses across all doctors and number n_i_ cases treated by doctor *i*. The z score is 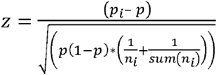. When sum(n_i_) is much larger than any n_i_ the z-score may be simplified to 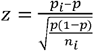. If z-scores are independent, the sum of *k* z-scores has mean 0 and standard deviation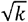.

Each doctor was grouped according to their grade to ensure fair comparisons. Any doctor with a z-score less than −3 or greater than +3 was considered an outlier. The doctors were selected from the extreme of both lowest and highest, moving towards the threshold of 3 standard deviations and were required to attend a face-to-face interview with a senior clinician and a senior member of Hospital Management, where any learning points, positive and negative, were shared. A written summary of the session was provided to the doctor and their educational supervisor. Following these interviews performance was reviewed after one month for the low performing end. If there was no significant improvement, the doctor was recalled to further interview and the process repeated. The Junior Doctor Clinical Dashboard was implemented in January 2013.

#### INTERVENTION 2 – ELECTRONIC PRESCRIBING: A MANDATORY ACTION FOR VTE ASSESSMENT AND PRESCRIBING

The PICS EHR was updated so that following completion of a VTE assessment module, an automatic prescription proposal was generated for either mechanical prophylaxis, pharmacological prophylaxis or both. The admitting doctor could authorise the VTE prophylaxis prescribing proposal or delete it but no further prescriptions could be added or modified to that patient record until a decision had been made on the VTE prophylaxis. Data on each step was captured in a structured form. This intervention was implemented in January 2014.

#### INTERVENTION 3 – AMENDING THE MANDATED PROFORMA FOLLOWING CLINICIAN FEEDBACK

The previous version of PICS allowed for the admitting clinician to choose ‘no reduced mobility’ early in the VTE prophylaxis proforma and thus quickly circumvent the automatic proposal for prescribing. After a risk review and feedback groups, it was apparent that in general, these decisions did not reflect actual VTE risk. A systems approach change to the VTE assessment form was made in consultation with prescribers, where the position of ‘no reduced mobility’ was moved from a check box at the top of the assessment to a much lower down part of the dropdown list of contra-indications, meaning the majority of the VTE assessment was completed prior to reaching this risk criteria. This change was made in October 2015.

#### FURTHER CHANGES TO PRESCRIBING PRACTICES

During the period of this analysis the Trust changed the brand and drug of Low Molecular Weight Heparin prescribed due to low stock levels from the drug company nationally. Once the stock levels were restored the Trust switched back to the original brand. Whilst this was not an intervention, it required the doctors to be trained on the new medication so the effect of these switches was measured as it occurred during this analysis.

For VTE, two-years of data were assessed prior to the implementation of the first intervention to gain baseline until October 2020 (a period of 9 years of prescribing practices)

### Statistical Analysis

Weekly values were used to avoid any day-of-the-week effect. The proportions were plotted against the weeks in chronological order. A probit transformation was applied to the proportions prior to analysis to achieve best fit of the assumptions required to then use a segmented linear regression model to determine whether there were significant step changes and/or significant changes in gradient at the times corresponding to the three interventions. Proportions at the start and end of each segment were estimated from this model and equivalent rates of change calculated by treating the change in proportions within each segment as linear. All analyses were performed using IBM SPSS Statistics Version 22.0 (Armonk, NY: IBM Corp.)

## Results

Data from 235,005 admissions were included in total, which represented all admitted adult patients during the study period. The number of patient admissions admitted in each timeframe, and the basic demographics of the patients are given in Table 1. There were no significant differences in patient characteristics during the study period.

**TABLE 1.** Demographics for patient admissions Values are counts (and percentages), except for age where they are medians and quartiles.

|  | Before intervention<br>(2 year run in period for data collection) | After introduction of Junior Doctor Clinical Dashboard | After introduction of mandatory VTE assessment and prescribing | After change in order of 'no reduced mobility' |
| --- | --- | --- | --- | --- |
| n | 31071 | 26260 | 39931 | 137743 |
| Female, n (%) | 14740 (47.4%) | 12675 (48.3%) | 19135 (47.9%) | 67340 (49.0%) |
| Age (years)* | 74 (59-86) | 73 (57-85) | 72 (57-84) | 68 (53-80) |

The number of prescribers during the period was 5503. Although the prescribers changed during this time (reflecting doctor’s rotations across regions during their postgraduate training) the seniority of doctor grades represented in the Trust did not alter over the study period, see Table 2.

**TABLE 2:** Prescriber Seniority Distribution of user type in the four main time intervals (values are percentage frequencies). There were no differences in prescriber seniority over the study period.

| Time interval | Consultant | Specialty grade | Core medical trainee | Foundation doctor | Staff Grade |
| --- | --- | --- | --- | --- | --- |
| Pre-intervention<br>(Jan 2011 – Dec 2012) | 19 | 32 | 17 | 29 | 4 |
| After intervention 1<br>(Jan 2013 – Dec 2013) | 18 | 33 | 15 | 27 | 7 |
| After intervention 2<br>(Jan 2014 – Sep 2015) | 17 | 34 | 14 | 27 | 8 |
| After intervention 3<br>(Oct 2015 – Oct 2020) | 16 | 33 | 11 | 32 | 8 |

Full guideline VTE compliance (risk assessment and correct action) within 24 hours of recommendation was 70.2% (230 out of 327 in an average week) in the period prior to any intervention (Jan 2011 – Dec 2012). It increased significantly to 77.2% (312 out of 404) in the period after the initiation of the Junior Doctor Clinical Dashboard (p<0.001).

There was another significant increase in VTE compliance to 94.7% (425 out of 449) in the period following the change in the EPS with mandatory VTE assessment (p<0.001). There was a small but significant decrease to 92.2% (479 out of 520) following the amendment to relocate “no reduced mobility” from the initial VTE checklist to a subsequent drop-down box (p<0.001). Weekly compliance is shown in Figure 1.

**Figure 1.**
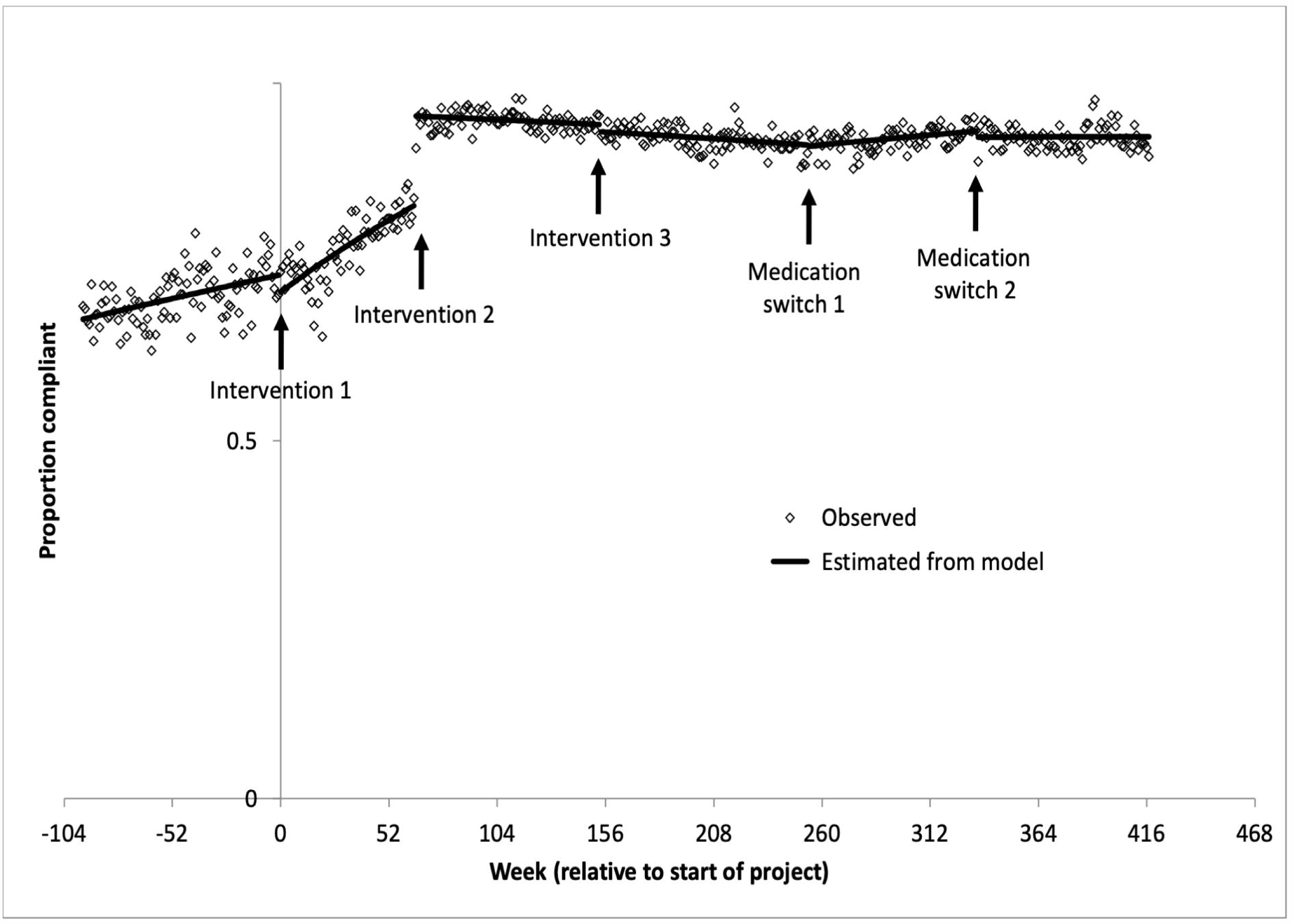
The proportion of patients who were fully guideline compliant over time. Graph showing the proportion of patients who were fully VTE guideline compliant, meaning they had both a risk assessment and then were either appropriately prescribed VTE prophylaxis or not, depending on that risk assessment. The regression lines are fitted over the time periods before intervention 1 (education/doctor’s dashboard), after intervention 2 (introduction of mandated VTE assessment action), after intervention 3 (change to PICS “no reduced mobility”) and Medication switch 1 (from enoxaparin to tinzaparin and then return to enoxaparin (Medication switch 2) until study end.

Table 3 summarised the VTE guideline compliance during six time periods. 1. Prior to any new intervention; 2. after the junior doctor dashboard interventions until the EHR mandate; 3. after the EHR mandate to the amendment of “no reduced mobility”; 4. after the amendment of the “no reduced mobility” option to the change from one LMWH to another brand; 5. to the use of this brand until a switch back to the original therapy; 6. from the reintroduction of the original therapy to study end.

**TABLE 3.** Observed full VTE guideline compliance over the study period Full VTE compliance is where a VTE risk assessment was completed and the correct action was taken. To be fully compliant, both VTE risk assessment and the correct action is needed. For example, non-compliance would be where a risk assessment was not completed, or a VTE assessment suggested LMWH was required and it was not prescribed or a VTE assessment suggested LMWH was not required (or contraindicated) and it was prescribed.

| Time interval | Length of interval (weeks) | Total number of admissions | Total number with full VTE guideline compliance within 24 hours | Compliance (%) |
| --- | --- | --- | --- | --- |
| 1. Baseline to intervention 1 (Junior doctor dashboard) | 95 | 31071 | 21809 | 70.2 |
| 2. Intervention 1 to intervention 2 (EPS system) | 65 | 26260 | 20264 | 77.2 |
| 3. Intervention 2 to intervention 3 (Change in risk order as presented) | 89 | 39931 | 37801 | 94.7 |
| 4. Intervention 3 to medication switch 1 (enoxaparin to tinzaparin) | 100 | 49931 | 46028 | 92.2 |
| 5. Medication switch 1 to medication switch 2 (tinzaparin to enoxaparin) | 81 | 45092 | 41583 | 92.2 |
| 6. Medication switch 2 to study end | 83 | 42234 | 38955 | 92.2 |

The estimates from the segmented linear regression model show that at time 0 (intervention 1: Junior doctor dashboard) compliance decreased from 73.2% to 70.7% but the rate of increase in compliance then changed from 3.4% per 52 weeks to 9.8% per 52 weeks. As a result, compliance increased to 82.9% by the time of the second intervention (the change in the EPS). After this intervention, there was a step increase to 95.4%. Subsequently, compliance remained relatively stable, although the third intervention (the change in order of “no reduced mobility”) produced a small step decrease from 94.2% to 93.2%. All the changes are described in Table 4.

**TABLE 4.** Estimated compliance from the segmented linear regression model and equivalent rates of change immediately before and after each intervention and medication switch Figures in parentheses for intervention 3 are p values if a slope change for intervention 3 is included in the model. As this was not significant, it was excluded and as a result the step change for intervention 3 became significant (the p value changing from 0.464 to 0.010). The final model includes five step changes and four slope changes.

| Intervention | Estimated compliance |  |  | Equivalent rate of change in compliance per 52 weeks |  |  |
| --- | --- | --- | --- | --- | --- | --- |
|  | Before | After | p value | Before | After | p value |
| 1. Junior doctor dashboard | 73.2 | 70.7 | 0.035 | 3.4 | 9.8 | <0.001 |
| 2. EPS implementation | 82.9 | 95.4 | <0.001 | 9.8 | -0.7 | <0.001 |
| 3. Change in order of risk criteria | 94.2 | 93.2 | 0.010<br>(0.464) | -0.7 | -1.0 | N/A<br>(0.085) |
| Medication switches |  |  |  |  |  |  |
| 1. Enoxaparin to Tinzaparin | 91.4 | 91.2 | <0.001 | -1.0 | 1.4 | <0.001 |
| 2. Tinzaparin back to Enoxaparin | 93.3 | 92.5 | 0.023 | 1.4 | 0.0 | 0.011 |

## Discussion

This study is the first to assess the cumulative impact of a series of interventions to improve full guideline compliant prescribing for VTE prophylaxis over a prolonged (nine-year) period, in a large NHS Trust which already offered a suite of educational tools such as lectures and videos and with high, and target compliant VTE assessment rates. This study highlights several important points.

First, that risk assessments do not automatically convert into an appropriate action following the assessment. Even after the introduction of the EPS system to mandate VTE risk assessment and appropriate prescribing, there was still a difference between completed risk assessments and prophylaxis prescribing. Altering reporting criteria to assess full guideline compliance may be a more effective means to improve patient safety.

Second, the interventions with demonstrable impact (the doctors dashboard clinic and rules based prescribing algorithms) require an EPS which supports dynamic evidence generation and application, enabling rapid learning and improvement based on data flowing from routine patient care. Both of these interventions were based upon the principles of a learning healthcare system, defined by the United States Institute of Medicine (now the National Academy of Medicine (NAM)) as systems where “science, informatics, incentives, and culture are aligned for continuous improvement and innovation, with best practices seamlessly embedded in the delivery process and new knowledge captured as an integral by-product of the delivery experience”^(21)^.

The most impactful intervention was a systems approach, with an EPS tool which mandated VTE assessment and prescribing as part of the admission process. These improvements were maintained at a higher level than seen following the individual feedback intervention, for the entire follow up period.

Our learning healthcare system included re-evaluating the PICS EPS to see where further improvements could be made. Feedback from prescribers and an assessment using the principles of human factors^(22)^ suggested that the placement of “no reduced mobility” at the top of the risk assessment algorithm potentially suggested that this was true for many patients and an audit of care records suggested that this was being erroneously applied in some instances. In light of this, the EPS was altered to place this option at the end of the list of contraindications, to ensure the prescriber considered co-morbidities and reason for admission prior to considering this option. This change did not have the impact expected on VTE compliance, and in fact contributed to a small, but significant, step decrease in full compliance. The reason for this is unclear and requires further study.

In this real-world study, there was a change in national LMWH availability, requiring a change in drug and prescribing rules (from enoxaparin to tinzaparin and then back to enoxaparin). These were introduced with traditional education but also necessitated a change in the EPS with a new series of prompts and rules. Despite the changes in prescribing practice, there was no significant change to full VTE guideline compliance, highlighting the resilience of the EPS systems-based approach.

This study did not assess why the systems approach was more effective than other interventions, but there are a number of potential reasons. As the Doctors Clinical Dashboard only identifies statistical outliers, only repeated failures to comply with guidelines will be identified. It is likely that many prescribing errors are made singularly and on an *ad hoc* basis, and these would not necessarily trigger a review. Educational and training events are one -off, and repetition in training has been shown to enhance performance^(23, 24)^. Healthcare is increasingly complex in terms of organisation and delivery^(25)^ and our ageing population often are multi-morbid and poly-medicated, making healthcare decisions more complex^(26)^. The complexity of healthcare and of patients might increase the potential for prescribing errors. A systems-based approach with prescribing support tools, that provides the same support for all prescribers on all occasions, is therefore more likely to impact on practice.

The current paper highlights the difference between VTE assessment compliance and full VTE guideline compliance (an assessment and appropriate prescribing action). While both are important, only the latter will reduce risk from HAT, but this information is not nationally collected or reported.

This study highlights the benefit of a paperless system, where real-time prescribing prompts can be given which account for clinical information, as opposed to static prompts, and where analysis includes all records overs a prolonged period. Some quality improvement papers in this field are based on standard audit procedures, where only a proportion of records are reviewed over a short period, leading to a significant risk of bias and making it unclear whether improvements were maintained^(27, 28)^.

Of note, the systems in place were unable to raise full VTE guideline compliance to 100%, and full VTE guideline compliance plateaued at approximately 92% (with risk assessment completion remaining >95% throughout). The reason for the small but important discordance in VTE risk assessment completion and subsequent correct action are unclear, but a further suite of electronically delivered tools are in development to determine if this can be improved.

This study has many strengths. It includes all patients within the hospital, and thus captures a high number of prescriber events in an unbiased manner. It also describes practices for a sustained period of time (nine years in total) which provides considerable reassurance that the changes in prescriber behaviour were sustained even as the workforce changed.

The paperless EHS deployed at UHB provides real-time, instantaneous feedback to all prescribers, highlighting the need for a VTE risk assessment, preventing further prescribing until this is completed and the suggested prescription is either approved or deleted. There are then further, automatically generated prompts every 24 hours to review the risk assessment. There are a number of reported interventions which provide retrospective feedback on VTE risk assessment and prescribing practices. These include a mandatory field within the electronic discharge system that record whether a VTE risk assessment on admission took place, the study of hospital coding on discharge or through audit ^(29)^. These provide the opportunity for learning but do not improve compliance or reduce risk for the patient included in the event. Other studies have suggested a VTE nurse specialist can provide real time feedback, reviewing notes in areas of low compliance and high risk^(30)^. This requires a significant workforce investment to operationalise a twenty-four hours a day, seven days a week service. An EPS solution is available to all, at all times.

This study also has limitations. All prescribing episodes were considered the same, while some guideline-discordant prescribing behaviour may be appropriate depending on the clinical circumstance. The study did not assess whether the improvement in prescribing practices benefited some patient groups or some specialities more than others. Nor did it assess whether the change in prescribing practices was associated with an improvement in patient outcomes (such as a reduction in VTE events) or reduced healthcare costs. Other studies have focused on in-patient HAT events and shown a reduction in the proportion of HAT attributable to inadequate thromboprophylaxis following an intervention with increased guideline compliance^(14)^ suggesting there would be significant clinical benefit from these interventions.

In summary, the use of mandatory assessment rules for VTE prophylaxis within an electronic prescribing system and continuous monitoring and feedback was successful in delivering and sustaining improved concordance between guidelines and prescribing practices in a large secondary and tertiary care hospital. Further work is required to determine whether these methods can be translated to other hospitals and whether these tools can be successfully used to improve performance in other areas. However, the significant and sustained impact demonstrated suggests this learning health systems approach, applied using routine clinical data to inform and refine practice, may demonstrate patient benefit across all areas of prescribing.

## Data Availability

To facilitate knowledge in this area, the anonymised participant data and a data dictionary defining each field will be available to others through application to PIONEER via the corresponding author.

## Author contribution

S Gallier, S Ball, T Pankhurst, M Berry, I Woolhouse, E Sapey designed the study, collated data, performed some analysis, wrote the manuscript. A Topham curated data and performed statistical analysis. P

Nightingale curated data and performed statistical analysis and helped write the manuscript. M Garrick supported data collection. All authors amended the manuscript and approved the final version.

## Acknowledgements

*This work was supported by PIONEER, the Health Data Research Hub in acute care and the HDR-UK Better Care programme. This work uses data provided by patients and collected by the NHS as part of their care and support. We would like to acknowledge the contribution of all staff, key workers, patients and the community who have supported our hospitals and the wider NHS at this time*

